# Prevalence and Outcomes in COVID-19 patients with AKI: A meta-analysis

**DOI:** 10.1101/2020.04.29.20079038

**Authors:** Zhixiang Mou, Tianjun Guan, Lan Chen

## Abstract

**Objectives:** The coronavirus strain first reported in December 2019 (COVID-19) has spread rapidly worldwide, posing a seriously risk to human health. This meta-analysis aims to shed much-needed light on the relationship between COVID-19 and AKI, and provide a stronger evidence base to support both further research and clinical application.

**Methods:** Two authors independently performed a literature search using PubMed, Web of Science, Embase, and Cochrane Library. Then the incidence of AKI, incidence of RRT required, the mortality rate with AKI and the risk of death with AKI during a COVID-19 infection were statistically analyzed using Open Meta-Analyst software, from which conclusions are derived.

**Results:** The incidence of AKI in hospitalized patients with the COVID-19 infection remains low, only about 3.8%; the in-hospital mortality rate with AKI in COVID-19 infected patients reaches up to 86.8%; the odds of death with AKI in COVID-19 infected patients is about 24.2 times higher than those without AKI.

**Conclusions:** The occurrence of AKI during a COVID-19 infection should be considered a strong red flag with regards to the patient’s risk of death. Additional studies are still required to support the conclusions derived herein and to explore the AKI mechanism during a COVID-19 infection.

## 1. Introduction

Starting in December 2019, a specific strain of the coronavirus disease was reported (named COVID-19). It is caused by the severe acute respiratory syndrome coronavirus 2 (SARS-CoV-2), exhibiting an unbelievable level of infectious distribution and resulting in high mortality rates worldwide [1]. According to the WHO, SARS-CoV-2 infections outside of China continues to increase rapidly; by April 20 2020, there have been 2,314,621 laboratory-confirmed cases and 157,847 confirmed deaths due to COVID-19 in over 150 different countries [2]. There are still no specific treatments or vaccines for COVID-19 at present, and as COVID-19 is a novel virus, developing our understanding of the virus is essential in establishing effective treatments.

The SARS-CoV-2 is identified as a positive-sense single-stranded RNA virus belonging to the β-coronavirus cluster, mainly causing community-acquired pneumonia in humans [1]. Fever and cough were the most common symptoms during infection [3]. Epidemiological data pertaining to COVID-19 thus far remains limited, with early studies focused on describing the clinical characteristics and severe cases. Recently, a growing body of evidence suggests that there is a common presence of acute kidney injury (AKI) in COVID-19 patients, including in patients who ultimately died [4-6]. Meanwhile, other studies [1,7] have cited AKI as only a rare incident during COVID-19 infections. More robust evidence is urgently needed to resolve clinical confusion and dispute. In addition, the effects of the COVID-19 infection on renal function still remain unclear and requires urgent exploration. This meta-analysis aims to investigate the authentic incidence rates of AKI during COVID-19 infections. By pooling the available data from relevant and valid publications, it aims to confirm whether COVID-19-related AKI has an impact on mortality or not.

## 2. Materials and Methods

### 2.1. Search Strategy

The protocol for this meta-analysis was registered with the International Prospective Register of Systematic Reviews (PROSPERO ID. CRD42020181188). A systematic literature search was performed using PubMed, Web of Science, Embase, and Cochrane Library from March 01 to May 04, 2020 (inclusive) to summarize the data of AKI on COVID-19 infected patients. The systematic literature search was not executed on any preprint databases to ensure the reliability of the final statistical results. Two authors (ZX.M. and L.C.) independently carried out systematic literature searches employing the terms “acute kidney” OR “acute renal” AND “coronavirus 2019” OR “novel coronavirus” OR “COVID 19” OR “2019 nCoV” OR “SARS 2” OR “severe acute respiratory syndrome coronavirus 2” OR “nCoV disease” OR “coronavirus disease 2019”. Each study was then evaluated for inclusion in or exclusion from this analysis. No language restrictions were applied. Publication date restrictions were applied from January 2020 up until May 04, 2020 (inclusive). A manual search for references cited in the articles found was also performed. The meta-analysis was conducted and handled according to the guidelines of the Preferred Reporting Items for Systematic Reviews and Meta-Analyses (PRISMA). (http://www.prismastatement.org/).

### 2.2. Inclusion/ Exclusion Criteria

For the meta-analysis presented herein and for our AKI definition, eligible studies reporting the AKI or AKI-related indicators of patients infected with COVID-19 were considered. Studies which met the Population, Interventions, Comparison and Outcomes (PICO) criteria were included in the meta-analysis.

All the studies selected fulfilled the following criteria, namely that original cohort studies provided: the necessary clinical characteristic data during COVID-19 infections; incidences of AKI requiring/not requiring renal replacement therapy (RRT) among COVID-19 infected patients; specific AKI-related indicators (hematuria, proteinuria, or serum creatinine) of COVID-19 infected patients are reported. All potentially eligible studies are considered for enrollment regardless of our AKI or RRT definition. All COVID-19 infected patients enrolled in this study were identified and confirmed as infected via nucleic acid detection by a laboratory.

Cases with a history of chronic kidney diseases pre-COVID-19 infections where it is impossible ascertain post-infection AKI in the original report were excluded; case or case series reports containing less than 10 patients were excluded; duplicate articles were excluded; reports from the National Health Commission were excluded in order to avoid repeated data.

### 2.3. Data Extraction and Study Quality

A data collecting spreadsheet (Excel: Microsoft Corporation, Redmond, WA) was created to analyze the following data from each included study: the first author, data collection period, region/hospital, number of patients, number of AKI/RRT patients, AKI-related indicators, AKI-related deaths. No attempts were made for specific or missing data from the authors. Due to the fact that some reports come from the same hospitals, and in order to prevent overlapping data, the duration and hospital were especially specified.

All included studies were assessed regarding their quality using the Newcastle-Ottawa Quality Assessment Scale (NOS) containing three aspects (selection, comparability and outcomes) and eight items [8]. Scores range from 0 to 9, where studies with a score ≧ 6 were considered as meeting adequate methodological quality.

### 2.4. AKI and RRT Definitions

RIFLE (the Risk of renal failure, Injury to the kidney, Failure of kidney function, Loss of kidney function and End stage kidney disease), AKIN (the Acute Kidney Injury Network), and KDIGO (the Kidney Disease Improving Global Outcomes) were the three major criteria to define AKI [9.10]. In this meta-analysis, AKI incidents were accepted regardless whether the original studies had been defined according to one of the guidance frameworks cited above; RRTs were accepted as the original studies have recorded the RRT used regardless of modality.

### 2.5. Statistical Analysis

Open Meta-Analyst software (for Windows 10 version; Brown University, Providence, USA) was utilized for the statistical analysis. In this analysis, random effects models and the DerSimonian-Laird method were applied to analyze the pooled incidence of AKI, the incidence of RRT required, the mortality rate with AKI (ratio variables), and the risk of death with AKI (dichotomous variables) during COVID-19 infection; the pooled odds ratio (ORs) was used to analyze the risk of death with AKI; all the corresponding 95% confidence intervals (CI) were evaluated; an *I*^2^ test was used to assess heterogeneity due to probability variance among observational studies; these results are graphically presented using forest plots. Statistically significant heterogeneity among studies was defined as χ^2^*P* value < 0.05 or *I*^2^ test > 50%; a *P* value of < 0.05 was considered statistically significant.

## 3. Results

### 3.1 Literature Search and Study Characteristics

The flowchart of the systematic review with selection process and reasons for exclusion are demonstrated in detail in Figure 1. A total of 61 records were identified from 4 databases (PubMed, *n*= 27; Web of Science, *n*= 7; Embase, *n*= 27; Cochrane Library, *n*= 0). After duplicates were excluded (32 records), 29 articles were screened through titles and abstracts manually, in order to establish eligibility. Then, in-vitro studies, animal studies, case reports, review articles, letters, comments, and erratums were excluded. The 18 remaining articles were full-text reviewed for eligibility. The cited references of the remaining articles were also screened through titles. The studies containing < 10 patients or insufficient clinical data and were subsequently excluded. Finally, 12 cohort studies involving 5,448 patients who fulfilled the pre-specified criteria were included in the meta-analysis (see Table 1). Four studies (33.3%) with a score ≧ 6 were considered to be of high quality according to the NOS criteria (Table 2) (as follow-up outcomes data were not available).

**Table 1.** Study characteristics

| Studies | Enrolled period | Region/Hospital | No. of patients | AKI | RRT | Proteinuria | Hematuria | AKI definition | Death with AKI |
| --- | --- | --- | --- | --- | --- | --- | --- | --- | --- |
| XL Zhang [11] | 17. Jan- 08. Feb | Zhe jiang | 645 | 2 | 0 | N/A | N/A | N/A | N/A |
| NS Chen [12] | 01. Jan- 20. Jan | Jin Yin-tan hosp | 99 | 3 | CRRT9 | N/A | N/A | N/A | N/A |
| T Chen [3] | 13. Jan- 12. Feb | Tong Ji hosp | 274 | 29 | CRRT3 | 100 | 84 | N/A | 28 |
| XB Yang [13] | 24. Dec- 26. Jan | Jin Yin-tan hosp | 52 (critical) | 15 | 9 | N/A | N/A | Scr increased | 12 |
| D Wang [14] | 01. Jan- 28. Jan | Zhong nan hosp | 138 | 5 | CRRT2 | N/A | N/A | KDIGO | N/A |
| CL Huang [15] | 16. Dec- 02. Jan | Jin Yin-tan hosp | 41 | 3 | CRRT3 | N/A | N/A | KDIGO | N/A |
| YC Cheng [6] | N/A | Tong Ji hosp | 701 | 36 | N/A | 194/442 | 118/ 442 | KDIGO | Hazard ratios |
| SB Shi [16] | 20. Jan- 10. Feb | Renmin hosp | 416 | 8 | CRRT2 | N/A | N/A | KDIGO | N/A |
| LW Wang [7] | 14. Jan- 13. Feb | Renmin hosp | 116 | 0 | N/A | 8 | N/A | KDIGO | N/A |
| Y Deng [17] | 01. Jan- 21. Feb | Hankou, Cai dian hosp | 225 | 20 | N/A | N/A | N/A | N/A | 20 |
| Richardson [18] | 01. Mar- 04. Apr | New York | 2634 | 523 | 81 | N/A | N/A | KDIGO | 347 |
| D Wang [19] | Jan-Feb | Zhong nan, Xishui hosp | 107 | 14 | N/A | N/A | N/A | KDIGO | 14 |

AKI= acute kidney injury; RRT= renal replacement therapy; CRRT= continuous renal replacement therapy; KDIGO= Kidney Disease Improving Global Outcomes; Scr= serum creatinine; hosp= hospital.

**Table 2.** Newcastle-Ottawa Scale for assessing the quality of cohort studies

| Studies | Representativeness of the exposed cohort | Selection of the non exposed cohort | Ascertainment of exposure | Demonstration that outcome of interest was not present at start of study | Comparability of cohorts on the basis of the design or analysis | Assessment of outcome | Was follow-up long enough for outcomes to occur | Adequacy of follow up of cohorts | scores |
| --- | --- | --- | --- | --- | --- | --- | --- | --- | --- |
| XL Zhang [11] | ☐ | ☐ | ☐ | ☐ | ☐ |  |  |  | 5 |
| NS Chen [12] | ☐ | ☐ | ☐ | ☐ | ☐ |  |  |  | 5 |
| T Chen [3] | ☐ | ☐ | ☐ | ☐ | ☐ |  |  |  | 5 |
| XB Yang [13] | ☐ | ☐ | ☐ | ☐ | ☐ |  |  |  | 5 |
| D Wang [14] | ☐ | ☐ | ☐ | ☐ | ☐ |  |  |  | 5 |
| CL Huang [15] | ☐ | ☐ | ☐ | ☐ | ☐ |  |  |  | 5 |
| YC Cheng [6] | ☐ | ☐ | ☐ | ☐ | ☐ | ☐ |  |  | 6 |
| SB Shi [16] | ☐ | ☐ | ☐ | ☐ | ☐ | ☐ |  |  | 6 |
| LW Wang [7] | ☐ | ☐ | ☐ | ☐ | ☐ | ☐ |  |  | 6 |
| Y Deng [17] | ☐ | ☐ | ☐ | ☐ | ☐ |  |  |  | 5 |
| Richardson [18] | ☐ | ☐ | ☐ | ☐ | ☐ | ☐ |  |  | 6 |
| D Wang [19] | ☐ | ☐ | ☐ | ☐ | ☐ |  |  |  | 5 |

Selection
Comparability
outcomes

**Fig 1.**
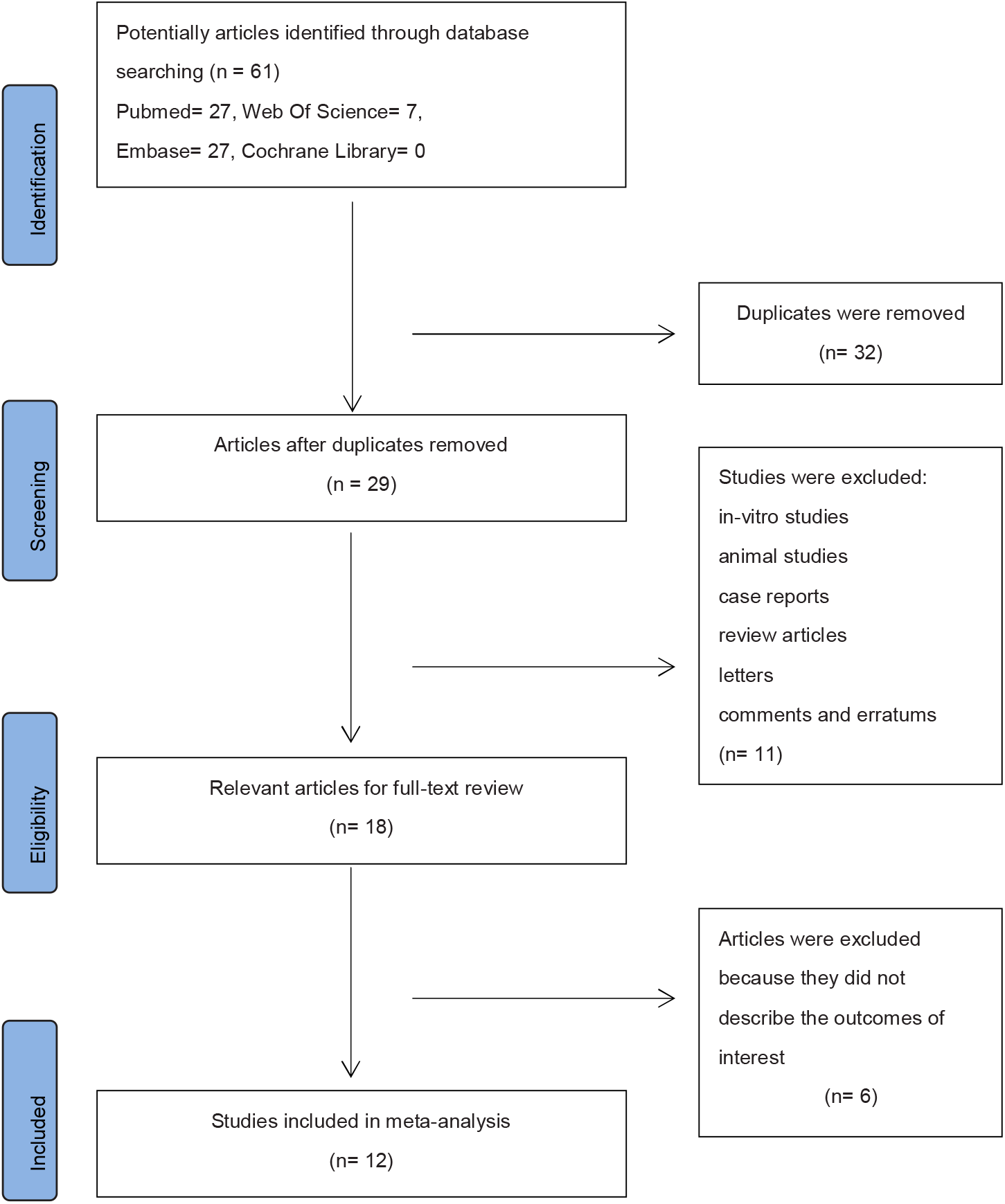
The flowchart for the systematic review.

### 3.2 Characteristics of Included Studies

The data collection period of enrolled studies is between December 16, 2019 and April 04, 2020. Seven studies (58.3%) reported the outcomes with clearly-defined AKI according to KDIGO. Eight studies (66.7%) reported RRT being required during COVID-19 infection and almost all of these used continuous renal replacement therapy (CRRT). Five studies (41.7%) clearly reported the outcomes with numbers of AKI-related deaths, while one study gave the hazard ratios (HR) between AKI and in-hospital deaths.

### 3.3 Incidence of AKI during COVID-19 Infection

Overall, 600 patients developed AKI during the COVID-19 infection, the pooled estimated incidence of AKI during COVID-19 infection being 3.8% (95% CI: 1.7% - 8.1%, *P* < 0.001, see Fig. 2), and a significant heterogeneity (*I*^*2*^ = 95.58%, χ^2^*P* < 0.001) was observed. Subgroup analysis was performed according to different AKI-defined criterion. The pooled estimated AKI incidences in the KDIGO subgroup analysis and none-defined (N/A) subgroup analysis are 4.6% (95% CI: 1.8% - 12.2%, *I*^*2*^ = 96.14%, χ^2^*P* < 0.001, *P* < 0.001) and 2.2% (95% CI: 0.4% - 13.9%, *I*^*2*^ = 91.08%, χ^2^*P* < 0.001, *P* < 0.001) respectively (see Fig. 2). There still remains significant heterogeneity following our subgroup analysis (data obtained by NS Chen [12] and XB Yang [13], duplicates from D Wang [14,19] were not calculated on two occasions).

**Fig 2.**
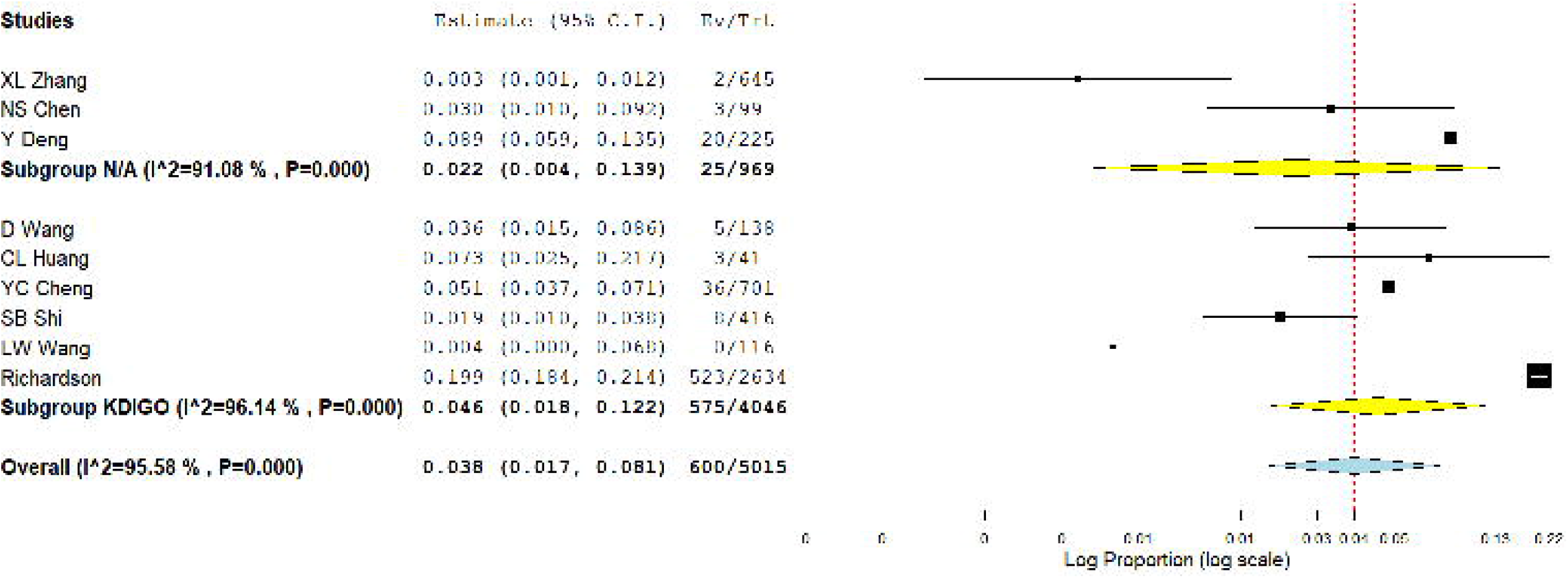
The overall incidence of AKI during COVID-19 infection.

### 3.4 Incidence of RRT Required during COVID-19 Infection

The following meta-analysis showed that 100 patients required RRT during the COVID-19 infection, the pooled estimated incidence of RRT required being 2.2% (95% CI: 1.0% - 4.6%, *P* < 0.001, see Figure 3), and a significant heterogeneity (*I*^*2*^ = 81.28%, χ^2^*P* < 0.001) is also observed among the eligible studies (data obtained by T Chen [3]and YC Cheng [6] were not calculated on two occasions).

**Fig 3.**
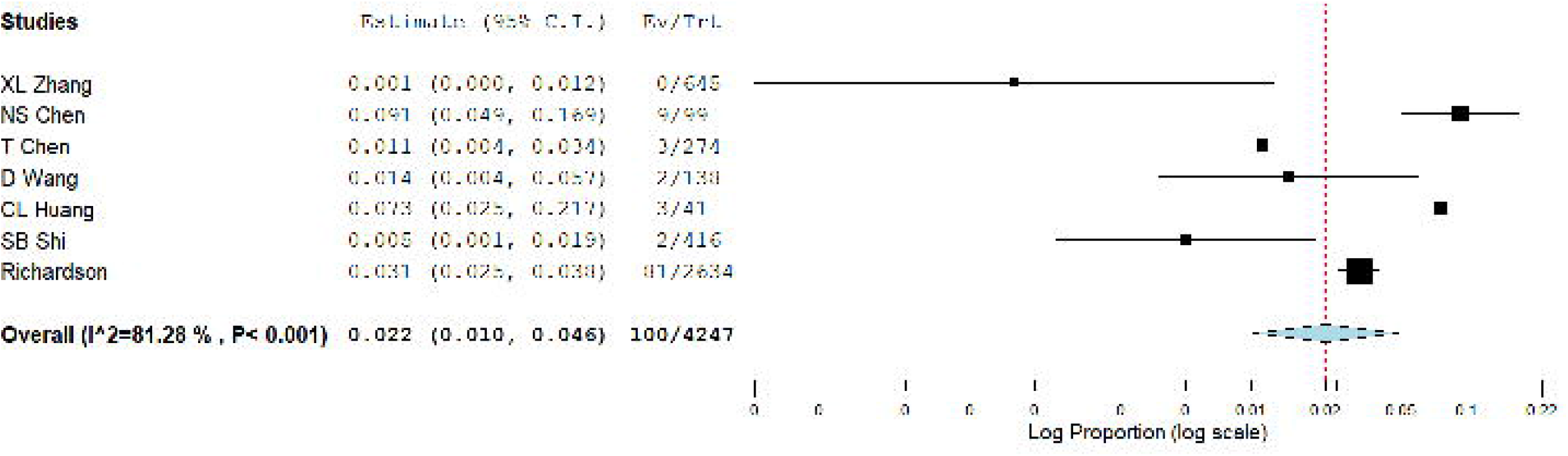
The overall incidence of RRT required during COVID-19 infection.

### 3.5 The Mortality Rate with AKI and the Risk of Death with AKI during COVID-19 Infection

The last series meta-analysis reveals that 421 patients died with AKI during their COVID-19 infection, the pooled estimated mortality rate with AKI being 86.8% (95% CI: 72.1% - 104.4%, *P* = 0.133, see Fig. 4a), with a significant heterogeneity also being observed among the eligible studies (*I*^*2*^ = 96.01%, χ^2^*P* < 0.001). The odds of death with AKI in COVID-19 infected patients is estimated to be 24.2 times higher than patients without AKI (OR, 24.2; 95% CI: 7.32 - 80.26, *P* < 0.001, *I*^2^ = 65.64%, χ^2^*P* = 0.02, see Fig. 4b), indicated that AKI was a strong risk factor for death. Subgroup analysis was performed due to different patients enrolled original studies. We could see that there was no heterogeneity (*I*^2^ = 0%, χ^2^*P* = 0.45, see Fig. 4b) observed in the three studies which conducted by enrolling all death and discharged patients, as well as being consistent with the overall results (OR, 18.6; 95% CI: 14.8 - 23.41, *P* < 0.001, see Fig. 4b). These results also pointed that further studies should carried out with clearly identifying variables patients.

**Fig 4.**
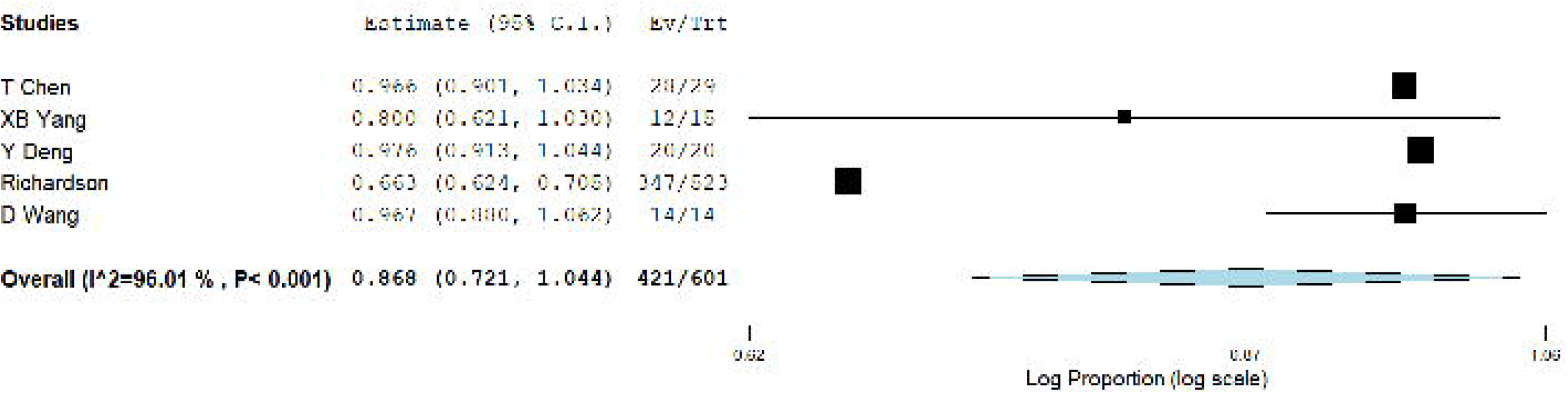
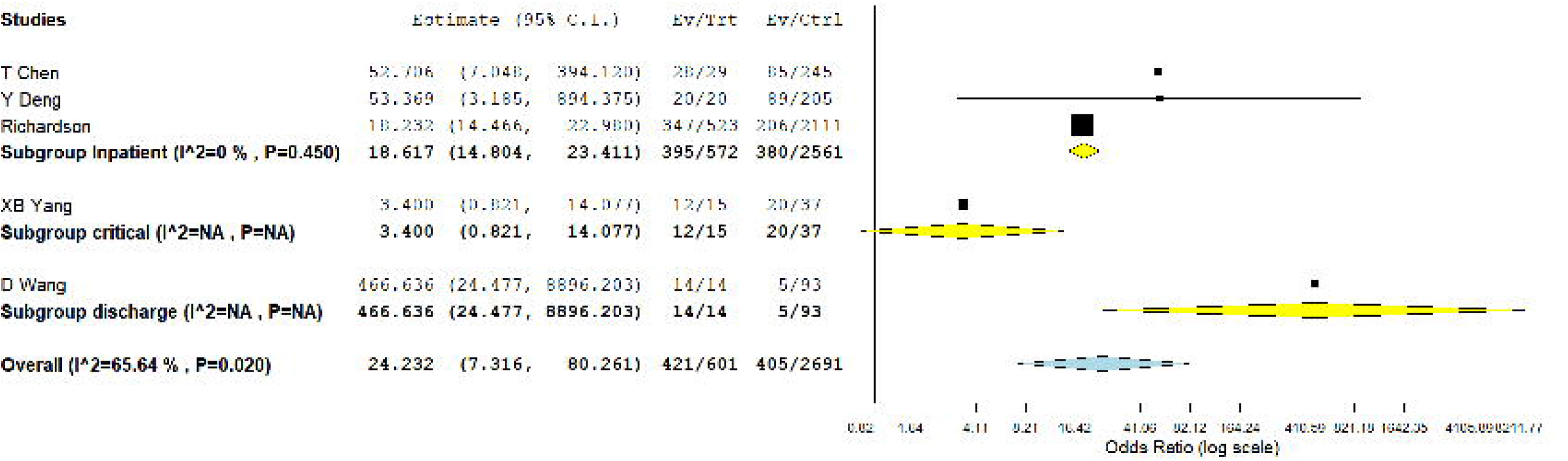
The dangers of AKI during COVID-19 infection. **a**. the overall Mortality rate with AKI. **b**. The risk of death with AKI during COVID-19 infection.

## 4. Discussion

This population-based meta-analysis provides much-needed evidence that the incidence of AKI in hospitalized patients with COVID-19 infections is low, at only about 3.8%. However, the appearance AKI during COVID-19 incidences infection should be recognized by clinical practitioners as a strong warning sign: the in-hospital mortality rate with AKI in COVID-19-infected patients is much greater, reaching as high as 86.8%; the odds of death with AKI in COVID-19-infected patients is extremely high, about 24.2 times higher than those without AKI.

The spike protein of SARS-CoV-2 binds to angiotensin converting enzyme 2 (ACE2), and would be activated and cleaved by cellular transmembrane serine proteases (TMPRSSs), leading to a release of fusion peptides, thus fusing with the host cells membrane [20]. Accordingly, the co-expression of ACE2 and TMPRSSs are key factors in determining the entrance of SARS-CoV-2 into host cells. One study, which is heavily based on analysis of normal kidney cells, indicates that ACE2 and TMPRSS genes exhibited high co-expression in podocytes and proximal straight tubule cells, which play critical roles in urine filtration, reabsorption and excretion [21]. According to the European Centre for Disease Prevention and Control (ECDC), about 80% of COVID-19 infection cases are mild in symptoms and outcomes, while 14% of patients experience severe infection, and 6% patients deteriorate to become critically ill [22]. As SARS-CoV-2 is predominantly first established via an induced respiratory tract infection, the duration and accumulation of SARS-CoV-2 in the circulatory system is likely to be a key mechanism by which to attack extrapulmonary organs, including the kidneys. Once extrapulmonary organs dysfunction manifests, such as in the kidneys, it can be inferred that a large number of viruses have replicated in the patient, with the effect that multiple organs are or will be attacked simultaneously. Moreover, a pathologic dysregulated host response with a hyper-inflammatory cytokine storm [23] combines to precipitate critical illness in patients, increasing the probability of death. Thus, the difference of infection duration, the various stages of the infection, and the inconsistent host conditions may together cause different AKI incidences and mortality rates during the COVID-19 pandemic. A time-dependent, anti-viral approach to treatment arguably needs to be focused upon, however, this is beyond the scope of this present meta-analysis. Nonetheless, renal function and the urine output of COVID-19 patients should be monitored early and frequently as a matter of course.

Recently, some studies have proposed a mechanism named “organ crosstalk”, implying that the impaired function of one organ is communicated to the dysfunction of other organs and is subsequently mediated among each other via complex mechanisms in critical illness [24,25]. AKI is one of typical examples involving multiple organ interactions [24,25]. AKI incidents associated with acute respiratory distress syndrome (ARDS) with variations of lung-kidney interactions have been identified [25]. It may act as an initiated incident or an aggravated factor via multiple mechanisms, including blood gas disturbances, renal congestion due to pulmonary hypertension, and hormone dysregulation [25]. In addition, mechanical ventilation-induced haemodynamics alternation and lung injury may amplify the conditions and exacerbate pre-existing or recently developed end-organ injury.^25^ As previously mentioned, no vaccine or specific anti-viral drugs for COVID-19 has yet been shown to be effective. At present, supportive treatments to ease the symptoms and to prevent multi-organ dysfunction in severe COVID-19-infected patients have been prescribed, which means executing interventions with the inevitable sacrifice of renal function (such as chloroquine phosphate). AKI might be considered as an inevitable clinical syndrome in these patients similar to like other critically ill patients, as over 50% of those would develop Stage 1 AKI at some point during their ICU course [26].

The present study provides a much-needed evidential basis for AKI incidence among COVID-19 patients, but has several limitations. First, there remains a limited amount of original studies (< 10) by which to evaluate the risk of death with AKI during COVID-19 infection. Nonetheless, considering the potential of this research in advancing our understanding of COVID-19 and in supplying medical practitioners with evidence-based recommendations, and in light of the current global pandemic emergency, the authors took the decision to present these findings forthwith. Second, there is a statistically significant heterogeneity in the meta-analysis for AKI incidence. Potential origins for significant heterogeneity may come from different COVID-19 infection stages. As performed via subgroup analysis according to AKI definitions, the results still reveal a significant heterogeneity. Future heterogeneity exploration must rely on the acquisition of more original clinical data. Finally, since the clinical data collection is based upon publications with limited availability, extra investigations could not be made but should be considered (infection duration, drugs used, geography, ethnicity, and age group in particular). Nevertheless, AKI incidence with COVID-19 infections must demand greater attention. Further studies are still urgently needed to support the conclusion herein and explore the AKI mechanism within a COVID-19 infection context.

Although the chance of AKI during COVID-19 is rare, it is a dangerous incident for these patients compare to those without AKI. Unfortunately, demographics of these patients in this meta-analysis are not possible to include. For example, we can not get the overall data of AKI in asymptomatic patients, who have neither fever nor respiratory tract symptoms, usually with good prognosis, yet the pathophysiological process of renal function changing even minor alternations of them should be worth for exploring.

Since mentioned the treatment of COVID-19 patients, there is also nearly no effective drugs directly targeting COVID-19 at present, which is similar to AKI. Prevalent prescription focused on antiviral activity including both the western medicine (lopinavir/ritonavir, remdesivir, arbidol, and chloroquine) and traditional Chinese antiviral medicine, that are equally missing effective and safe evidence from multi-center, randomized controlled and double-blind studies. Usually, it would take quite a long time from *in vitro* experiments to actual clinical application for any medicine. However, the nephrotoxicity of the drugs above-mentioned are already proved for many years. In addition, for critical illness patients infected by COVID-19, strengthened pharmaceutical care or antiviral treatment might be inevitable thereby often conducted. As there are lack of real effective drug to prevent or cure kidney damage during the early stage of this complex lung-kidney interactions process, renal function/urine output frequently monitored should be carried out, together to apply drugs with low nephrotoxicity where possible, accordingly to reduce potential or actual drug problems once virus detection verified, based on the conclusions of this meta-analysis.

The reason of establishment this meta-analysis is not only to generalize the dangerous of AKI during COVID-19, but also to provide suggestions for making treatment decisions. The antiviral effect of COVID-19 medicine should not be the only judging criteria. According to the complex mechanisms of AKI during COVID-19 and its relationship to the risk of death, further exploring of new COVID-19 targeting drugs could not only limited to the antiviral effect, the anti-inflammatory (anti-IL-6, anti-TNF-α, and so forth.) ability should also be one of consideration for the new drugs development. The second treatment solution may be powerful antiviral drugs combined with anti-inflammatory drugs especially for critical patients, due to the possibility of those medicine playing important roles in neutralizing the impact of cytokine storm, reducing the incidence rate of AKI, finally decreasing and the mortality rate of critical patients.

In conclusion, the occurrence of AKI during a COVID-19 infection should be paid greater attention, and should be considered a strong red flag with regards to the patient’ s risk of death. Additional studies are still required to support the conclusions derived herein and to explore the AKI mechanism during a COVID-19 infection. The following fields should be emphasized for the reduction of mortality related AKI during COVID-19: (1) Strengthen epidemiological research, especially the prevention and control of critical cases with AKI; the renal function of asymptomatic and mild cases should be frequently monitored, carried out blood purification treatment if necessary; (2) Conduct in-depth research on mechanisms of renal injury during COVID-19 attack (various signal pathways/virus proteins) for designing of blocking drugs; (3) Explore the combined application of antiviral and anti-inflammatory drugs in the design of clinical trials for COVID-19, further study is needed on the standard application, the evaluation for the curative effect, and the integration of them together. Finally to ameliorate the effectiveness of treatment, as well as to achieve the goal of reducing mortality patients infected with COVID-19.

## Data Availability

All data generated or analyzed during this study are included in this article.

## Declarations

### Acknowledgements

Not applicable.

### Authors’ contributions

C.Lan had the idea for the study and formulated its design, having had full access to all data in the study and takes responsibility for the integrity of the data and the accuracy of the data analysis. Z.X. Mou and T.J. Guan contributed to data acquisition and the writing of the report. Z.X. Mou contributed to critical revisions of the report. Z.X. Mou contributed to the statistical analysis. All authors commented on previous versions of the manuscript. All authors read and approved the final manuscript.

### Data availability

Not applicable.

### Consent to publish

Not applicable.

